# An Analysis Between Trump’s Presidency and Health-Seeking Behaviors of African Americans in the United States

**DOI:** 10.1101/2022.02.17.22271104

**Authors:** Adeola A. Animasahun, Ashley M. Nunez, Rachelle F. Jean

## Abstract

**Background:** Slavery legally ended over 150 years ago, yet African Americans are still oppressed. The lingering effects of systemic and institutional racism are still present in all walks of life, especially in healthcare. As a result, Black patients have historically been less likely to seek preventative care and subsequently, reported lower health outcomes compared to their white counterparts. The election of Donald Trump in 2016 forced a shift in that narrative. This study aimed to investigate whether there was any correlation between changing racial preferences fueled by Trump’s racist rhetoric and the health-seeking behaviors of Black patients. Through this, we explored if the number of Black patients that reported not having a primary care physician has changed, and how online search history trends researching Black physicians have also changed.

**Methods:** This study utilized datasets from the Behavioral Risk Factor Surveillance System (BRFSS) and Google Trends. Pearson’s correlation testing was run to establish any correlation between the number of google searches and positive health-seeking behaviors.

**Results:** GoogleTrends data does support an increased popularity for the search term “black doctor near me” over the years of 2015-2018, supporting our hypothesis. Multiple logistic regression analysis was performed to determine if race, among other variables, was a significant predictor for our predetermined health indicators. The results showed race was significant in nine out of our eleven health indicators.

**Discussion:** Despite decades of work to minimize healthcare disparities, this study has demonstrated how much more still needs to be done. It has shown how the intersection of seemingly unrelated issues, such as politics and health, as described in this study, can drastically impact health outcomes. This study highlights the importance of targeted and equitable programming to ensure quality care for all, that can withstand political and social pressures.

## INTRODUCTION

In recent years, there has been an unprecedented focus on racial inequalities, racially-based brutality, and a heightened sense of racial divide within the country^1^. The focus of racial inequalities directed towards the Black community has specifically been highlighted, and the Black community has attempted to push back against instances of disparity and racism as can be seen through the large activity and support of the Black Lives Matter movements. According to FBI Uniform Crime Reporting, hate crimes can be defined as “a committed criminal offense which is motivated, in whole or in part, by the offender’s bias(es) against a race, religion, disability, sexual orientation, ethnicity, gender, or a gender identity”^2^. Recent research has identified from FBI statistics a significant increase in hate crimes across the United States since 2016^1^. It is important to acknowledge that a number of Black individuals in the United States have been negatively affected by instances of bias: In 2020, 65% of Black men and 59% of Black women believe that it is a bad time to be Black in America^3^ and that 65% of Black adults report that individual acts of discrimination have been a personal obstacle^4^. Healthcare is not exempt from disparity and bias. It has been established that distrust of physicians is prevalent among Black patients and has been correlated to the instance of not having a regular health care provider^5^. In light of this information, Black individuals are less likely to have a regular health care provider due to high distrust of physicians^3,6^, and visit office-based physicians at low rates^3^ despite reporting poor health status and outcomes^6^. This is a problem in public health due to failure to receive preventative care or medical treatment in general, leaving chronic health issues unmanaged. This is especially important among patients given their higher rates of mortality and poorer outcomes^5^. For years, Black patients have reported feeling ignored by physicians; previous research discusses how prejudice exists in medicine, with physicians treating pain management differently depending on race, due to physician implicit bias and beliefs^4^. In light of this information, recent studies show that Black patients tend to have better health outcomes and place more trust in their physicians when their physicians are of the same race^7,8^. The study aimed to further evaluate how racism and disparity, quantified by the instances of anti-black hate crime, transfers to the world of medicine. It is important to distinguish if recent events have further pushed Black patients away from seeking care. It is important to also identify any other changes in health-seeking behaviors—such as if the desire to seek treatment from physicians of the same race has increased—in order to understand how to provide better care to Black patients.

The increase of racial tensions can be attributed to the divisive and racist rhetoric, incited by Trump’s presidency^8^. Since his election in 2016, there has been approximately a 20% rise in hate crime incidents^9^. Trump’s presidency, in conjunction with the rise in hate crimes, has continued to make Black patients less likely to seek preventative care despite Black patients being more likely to have poorer health outcomes than their white counterparts. For decades, healthcare disparities have been fueled by a combination of factors, including socioeconomic status, access to healthcare, along with medical mistrust, perceived discrimination, and many others^6^. Black patients often turned away from preventative care due to perceived racial discrimination, which past researchers have associated with a range of adverse health outcomes^10^. Another study demonstrated how racial discrimination exists by using an example of non-cancer pain management^11^. They demonstrated that Black patients were less likely to receive more aggressive treatment compared to their white counterparts presenting with the same diagnosis and level of pain^11^. Other research then identified the presence of implicit biases among physicians and how those with a pro-white bias ultimately were more dominant and verbal towards their Black patients, making them feel less respected and listened to, and thus had less agency in their treatment plan^12^. While factors contributing to these disparities have been identified, there is very little research describing how racially charged events, such as Trump’s presidency, has increased mistrust in the healthcare system, mainly for Black patients^6^. A poll was performed in January 2020, asking Black adults in the United States what they thought of the current state of affairs, and reported that 83% of respondents believed racism had become a bigger issue in the United States since Trump started his presidency^13^. Some scholars even identified a strong correlation between cities where Trump held rallies in 2016 and a significant rise in hate crimes^2,8,9^. These same cities, such as Atlanta and Fort Worth, Texas, also saw a significant increase in the number of google searches of a “black doctor near me”. Certain actions by the Trump administration such as trying to repeal Obamacare, or at least restrict some of its functions, along with its initial inaction with the COVID-19 pandemic, ultimately led to millions of Americans losing health insurance, all of which has caused minority populations to be disproportionately affected by the virus^14,15^. In New York, Black communities were severely impacted during the presidency of former President Trump^16^. Many Black patients lacked trust in their non-Black doctors because of prior studies of racial biases against Black patients. The Trump Administration impacted this group additionally, by proceeding with the attempt to remove the Affordable Care Act, also known as Obama Care^17^. This Act was set in place by former President Obama, to give millions of Americans the opportunity to be able to afford health insurance^17^. When the Trump Administration attempted to remove this healthcare proxy, it risked leaving millions of Americans without health insurance^17^. Such an action would negatively impact many low-income and minority groups at disproportionate rates, especially Black patients, leaving these individuals without coverage^17^. As previously mentioned, a number of studies have shown that prejudiced remarks made by President Trump, led to the harboring of negative remarks and rhetoric aimed at minority groups, such as Black individuals^8,12,18^. Numerous occurrences of targeted brutality, injustices, and discrimination have disproportionately impacted the black community, further diminishing the trust that Black individuals have in their communities^4^. The change in racial attitudes that are seen today has forced Black people to become more intentional about the care they seek. Black patients have become more invested in concordance within their physician-patient encounters so that there is some shared identity between them, which ultimately “improves medical compliance and quality of care^7^.”

This study aimed to determine whether the increasing racial tensions from 2015-2018 impacted whether Black patients reported having a primary care doctor, and to determine whether they influenced online physician search strategies. The first hypothesis for this study was that there would be an increase in search term hits for “a black doctor near me” from 2015-2018. The second hypothesis was that there would be a correlation between online Google searches for Black physicians and positive health seeking behaviors. The third hypothesis was that race would be a strong predictor of various health indicators.

## METHODS

### Participants

The BRFSS dataset utilized different survey questions every year and each state chose which additional health modules, if any, they also wanted to include for their particular survey, with their results in several data files. To keep the health indicators consistent within the target population and easier to analyze, the target population was chosen from states listed on the BRFSS website as having the same health modules being analyzed from 2015-2018, contained within one datafile “LLCP” (Combined Land Line and Cell Phone data). Thus, the target population for this study consisted of all adult White and African American individuals living in two different states with opposing political views, the District of Columbia and Louisiana. These two areas were chosen specifically because they utilized the same questionnaire with the same health indicators in the original BRFSS dataset in all four study years. Furthermore, these areas also had opposing political views with Trump having a much higher approval rating in Louisiana and DC having a lower approval rating for Trump^23^. The source population will include individuals who identified as Black, non-Hispanic, and individuals who identified as White, non-Hispanic to the BRFSS questionnaire in DC and Louisiana, from 2015 to 2018. Respondents that reported being mixed raced were excluded. Responses from U.S. territories Guam and Puerto Rico were not included. The study population size (n) meeting the above criteria was 32,195 across all four years, 2015-2018^18^.

### Procedures

Secondary data analysis in this study was drawn from the BRFSS and Google Trends datasets. Data from Google Trends was used to observe changes in the popularity of searches for “black doctors near me” over the years 2015-2018. Google trends data on the search term’s popularity served as one way for measuring health-seeking behaviors. A correlational approach was used to identify any trends between the rise of online Google searches and health-seeking behaviors in finding physicians along with overall health. The BRFSS dataset was used to observe changes in the number of Black patients who reported not having a primary care physician. The outcome was measured as the number of Black individuals who reported not having a personal health provider per 100,000 individuals per comparison year per state. The data from the study has been de-identified so there is minimal risk to participants. This study underwent review by the Geisinger IRB Committee at the start of the CHR 2 course and was exempt.

### Data Analysis

A table was generated utilizing the BRFSS data on responses from Black participants on the health indicators listed in Table 1. The second table generated showed the number of relevant Google Trends search terms by year in the United States. The study analyzed any overall changes, such as overall decline or increase in the number of Black patients without PCPs and any changes in overall health and other health indicators between the years 2015-2018. Data that met the study’s timeline and criteria were selected to compare and relate observed changes over time. A logistic regression analysis was performed to determine any effect of race on various health indicators from 2015-2018.

**Table 1.** Health Indicators Evaluated and Parameters Used for Logistic Regression Analysis. List of all eleven health indicators utilized for the logistic regression analysis. Also included are the expanded health variable questions, reference value, and comparison value. The results from the logistic regression all stem from the comparison category.

| Table 1. Health Indicators Evaluated and Parameters Used for Logistic Regression Analysis |  |  |  |
| --- | --- | --- | --- |
| Variable | Question | Reference Value | Comparison |
| X_RACE | Race/ethnicity categories | White only, non-Hispanic | Black only, non-Hispanic |
| ADDEPEV2 | (Ever told) you have a depressive disorder (including depression, major depression, dysthymia, or minor depression)? | No | Yes |
| CHECKUP1 | About how long has it been since you last visited a doctor for a routine checkup? | Over a year ago | Once within the past year |
| MEDCOST | Was there a time in the past 12 months when you needed to see a doctor but could not because of cost | No | Yes |
| MENTHLTH | Now thinking about your mental health, which includes stress, depression, and problems with emotions, for how many days during the past 30 days was your mental health not good? | Less than 14 days | 14 days or greater |
| PERSDOC2 | Do you have one person you think of as your personal doctor or health care provider? | No or unsure | At least 1 |
| POORHLTH | During the past 30 days, for about how many days did poor physical or mental health keep you from doing your usual activities, such as self-care, work or recreation? | Less than 14 days | 14 days or greater |
| X_HCVU651 | Respondents aged 18-64 who have any form of health care coverage | Not covered | Covered |
| X_RFBMI5 | Adults who have a body mass index greater than 25 (Overweight or Obese) | No - BMI less than 25 but greater than 12 | Yes - BMI greater than 25 |
| X_RFHLTH | Adults with good or better health | Fair or Poor Health | Good or Better Health |
| X_RFSMOK3 | Adults who are current smokers | No - not a current smoker | Yes - a current smoker |
| X_TOTINDA | Adults who reported doing physical activity or exercise during the past 30 days other than their regular job | No - no physical activity or exercise in the last 30 days | Yes - had physical activity or exercise |

We used GraphPad Prism version 9.0.0 for MacOS, R 4.1.2 (MacOS), Excel (MacOS), Numbers (MacOS) and GoogleSheets to perform our statistical analyses and generate our figures.

We highlighted the change in Google search trends through GoogleTrends and illustrated the trend as shown in Figure 1.

**Figure 1.**
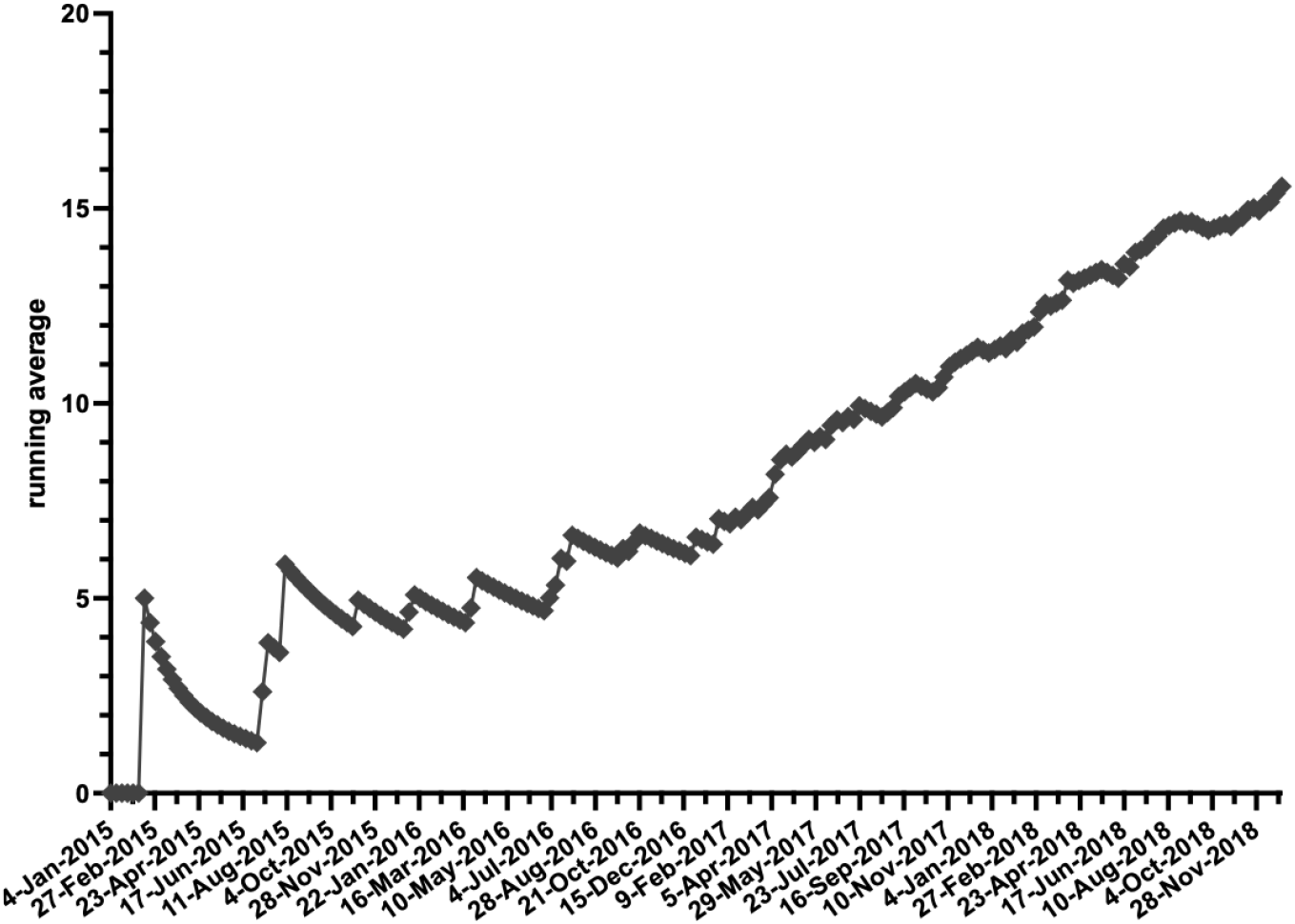
Increase in the number of “black doctor near me” searches in the United States over the past five-years with GoogleSheets^22^.

## RESULTS

A total of 32,195 responses from individuals that participated in the BRFSS questionnaire across DC and Louisiana from the years of 2015-2018 and fitting the inclusion criteria were retained. 7,890 responses from 2015, 8,139 responses from 2016, 7,761 responses from 2017 and 8,405 responses from 2018, make up the total sample size of 32,195. All responses were from individuals that lived in DC and Louisiana, identifying as “Black.”

Google Trends was utilized in order to monitor the popularity searches for the term “black doctor near me”, over time. Since Google Trends data represents search interest relative to the highest point on the chart for the given region and time, a time period between January 1, 2015 and December 31, 2018 was selected. In the selected time frame, a value for popularity in the search term is given each week. The popularity for the search term steadily increased between 2015 and 2018 as seen in the running average (Figure 1). The average popularity for each year was calculated and plotted which showed significant increase from 2015 to 2018 (Figure 2).

**Figure 2.**
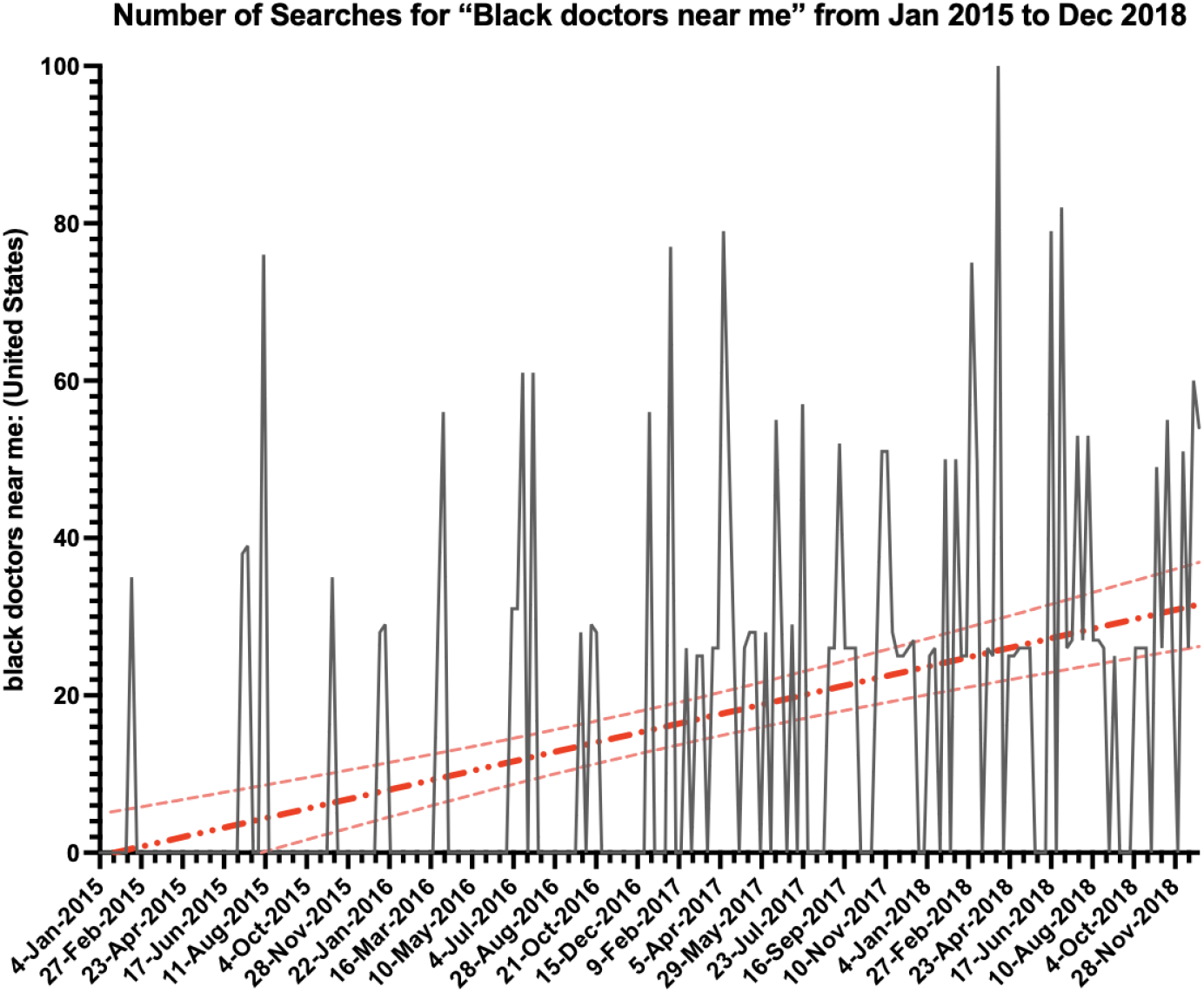
Average Searches for Black Doctors Near Me. The average popularity for the search term “black doctor near me” for each year was calculated, plotted, and fitted to a regression line to show significant increase (F=46.25, p=<0.0001) from 2015 to 2018.

**Figure 3.**
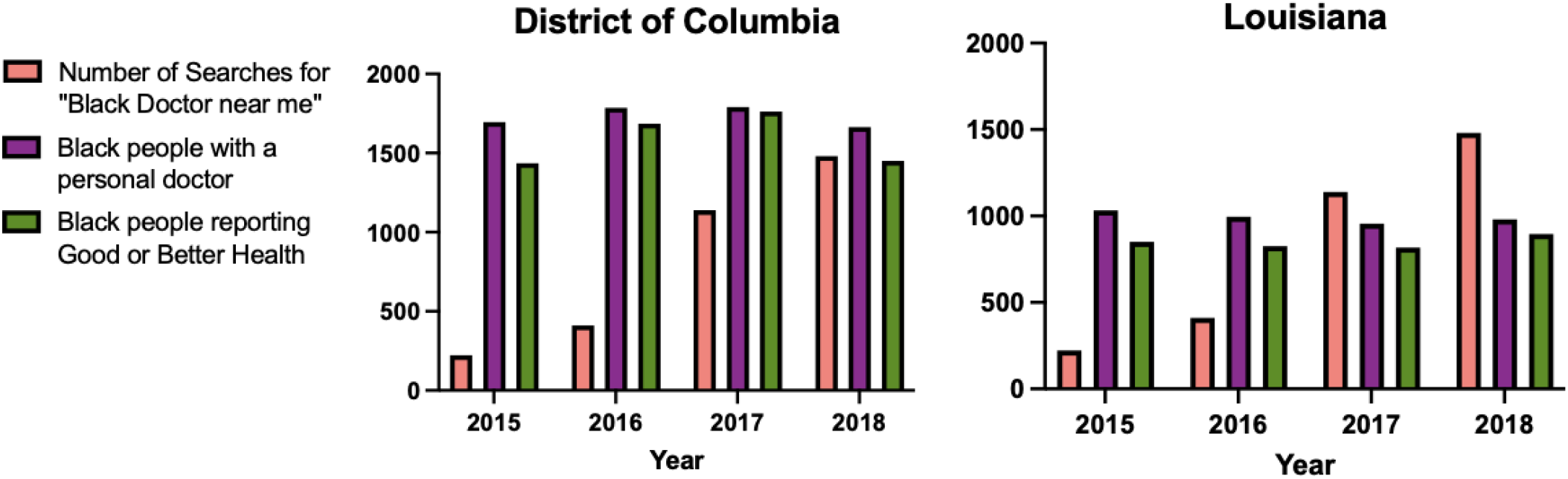
Data for the number of online Google searches from 2015-2018 was compared with the number of Black people with a personal doctor and the number of Black people reporting good or better health, in DC (A) and Louisiana (B).

Data for the average number of Black Americans with primary care providers for the years of 2015-2018 was compared with the number of online Google searches and their self-reported status of their overall health graphically. Nationally, as the google searches increased, more Black people in DC reported good or better health and had a personal doctor, however the same trend was not seen in Louisiana.

The number of all Black Americans in each state who reported having a Primary Care Physician for each year (2015-2018), among several other health indicators, were compared using logistic regression analysis. Regression testing suggested Black people were 0.75 times less likely than whites to have a personal doctor in Louisiana, which was statistically significant in 2015, 2016, and 2018 (p = 0.00271, 0.036, 0.014 respectively). In DC, Black people were actually 1.3 times more likely to report having a personal doctor than white people, although this was only significant in 2016 and 2018 (p = 0.0228 and 0.021 respectively) (Figure 4). Outside of having a personal doctor, the odds of reporting certain health indicators followed a similar trend in both DC and Louisiana, although the odds were higher in DC due to a higher response rate from Black residents.

**Figure 4.**
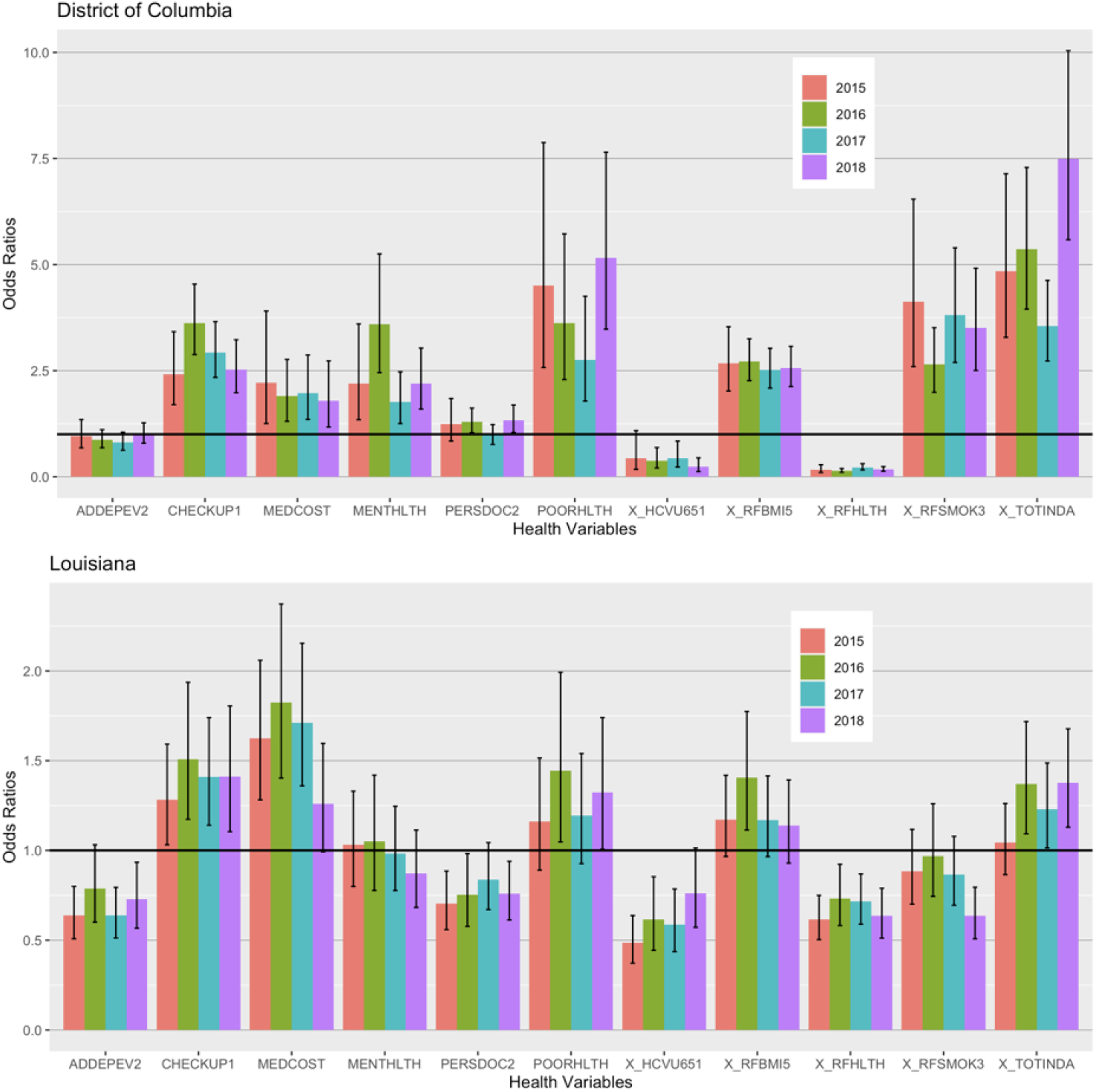
Odds Ratios of Eleven Health Indicators from DC and Louisiana between 2015-2018. Graph depicts the odds of Black people having the specified health indicator, compared to their White counterparts. Error bars illustrate the 95% confidence interval for each odds ratio. A horizontal line was also included at y=1 to highlight the point where there was no significant difference between Black and white people for that particular health indicator. Any Odds Ratio, including the 95% CI that crosses the y=1 line, was not significant, and anything below or above that line was significant. From left to right, “ADDEPEV2” - Ever told you had a depressive disorder? (Yes); “CHECKUP1” - About how long has it been since your last routine checkup? (within the past 12 months); “MEDCOST” - Was there a time in the past twelve months when you needed to see a doctor but could not due to cost? (Yes); “MENTHLTH” - How many days in the past month was your mental health not good? (14 days or greater); “PERSDOC2” - Is there someone you think of as your personal care doctor? (Yes, at least one); “POORHLTH” - How many days within the past month did poor physical or mental health prevent you from doing your normal activities? (14 days or greater); “X_HCVU651” - Respondents aged 18-64 with any form of healthcare (Covered); “X_RFBMI5” - Adults with a BMI over 25 (Yes, greater than 25 - overweight or obese); “X_RFHLTH” - Adults who report good or better health. (Yes); “X_RFSMOK3” - Adults who are current smokers. (Yes); “X_TOTINDA” - Adults who report doing physical activity outside of work. (Yes).

## DISCUSSION

Our data suggests that there were significant changes observed in negative health-seeking behaviors in Black Americans between the years of 2015-2018. Our data also suggests that there was no correlation found in trends in online Google searches and the average number of Black Americans reporting having a primary care provider, or their overall health, contrary to our hypotheses. GoogleTrends data does however support an increased popularity for the search term “black doctor near me” over the years of 2015-2018, supporting our hypothesis. This suggests that between the years of 2015-2018, individuals within the United States, for unknown reasons, were particularly interested in locating specifically Black physicians for care. Although there was no clear relationship between GoogleTrends data and the number of Black people with a personal doctor, race was a clear and significant predictor for the majority of the health indicators, which confirms previous research describing differences in care based on race^3,5,9.11^. Trump’s presidency was used as context to explain these results simply because of the racist rhetoric his term promoted^4,7^. As a result, Black people were more inclined to see a doctor of similar background (Figure 2). Although there were significant increases in these searches, the same trend was not seen in the number of Black people reporting a personal doctor, which is attributed to insurance coverage, cost, transportation, among many other factors^14^. When the Affordable Care Act was first enacted by President Obama, it aimed to minimize the large disparities present in healthcare coverage among the different races and ethnicities. However once Trump took office, some of his policies negatively targeted the Affordable Care Act causing many to lose coverage and prevented many more from getting coverage^13,16,24^. The effects of which were quite immediate with Black people being significantly less likely to have healthcare coverage in a more progressive area (DC) and even in a more conservateive area (Louisiana) (Figure 4).

## LIMITATIONS

There are a number of limitations that have been identified within the study model outlined here. Limitations in using Google Trends data have been identified, such that it is not the only search mechanism patients used to locate a Black healthcare provider. Searches for “a black doctor near me” might be made by individuals that do not align with the population meeting the inclusion criteria for this study. In addition, Google Trends data cannot determine each individual’s reasoning behind completing the search. Additionally, the BRFSS dataset does not gather information from every resident of each state, which may alter the accuracy of representation for results, however BRFSS has incorporated raking to weight data according to known proportions of race and age in a population. Lastly, by only evaluating two states in the US, it may have made my data less generalizable to all Black people in the United States.

## FUTURE RESEARCH AND IMPLICATIONS

The basis of this and similar studies explore the implications of racially motivated biases across the United States on medical mistrust in Black patients^3, 5^. Further continuation of this study would be necessary to target specific demographics within the Black community that have been affected the most by unconscious bias. The logistic regression analysis performed earlier can be expanded to include other predictors besides race such as age, education, income, employment, metropolitan status, etc, along with any interactions between them. Studies^6,9,10^ such as the one presented here could allow the development of community level interventions by focusing on building trust and lessening the experience of discrimination during physician-patient encounters. One way to improve this is to suggest that physicians get more involved in the communities that they serve; to learn about the disparities experienced by Black patients and to build rapport with the community, as suggested by previous polling^3^. Another possible intervention would include creating community programs to educate Black patients and the community about resources on local Black providers that they are actually comfortable with. Programs optimized for cell phones could also be installed that teach Black patients how to be an active participant in the management of their care, in order to harbor positive health seeking behaviors. Future quantitative and qualitative studies would benefit by gathering primary data from participants who directly experienced instances of bias in the medical setting. Data from participants who have personally experienced bias in patient-physician interactions would be best to evaluate causative implications of bias on health-seeking behaviors, and effects on patient’s trust in physicians. Extensive intervention is required to effectively minimize the effects of unconscious bias and blatant racism in our fractured healthcare system in order to ensure quality care for all.

## Data Availability

All data produced in the present work are contained in the manuscript

## Acknowledgements

This research paper would not have been possible without the amazing support of my supervisor, Dr. Brian Piper. His vast knowledge and attention to detail has been my motivation to keep my work on track despite all of the hurdles we jumped through, to ultimately reaching a final draft. Dr. Reema Persad-Clem not only has been a great advisor but also helped my team and I develop a thoughtful research question that was near and dear to our hearts. Iris Johnston was a great resource for my team in our initial database search.

